# Association of neurotransmitter pathway polygenic risk with specific symptom profiles in psychosis

**DOI:** 10.1101/2023.05.24.23290465

**Authors:** Tracy L. Warren, Justin D. Tubbs, Tyler A. Lesh, Mylena B. Corona, Sarvenaz Pakzad, Marina Albuquerque, Praveena Singh, Vanessa Zarubin, Sarah Morse, Pak Chung Sham, Cameron S. Carter, Alex S. Nord

## Abstract

A primary goal of psychiatry is to better understand the pathways that link genetic risk to psychiatric symptoms. Here, we tested association of diagnosis and endophenotypes with overall and neurotransmitter pathway-specific polygenic risk in patients with early-stage psychosis. Subjects included 206 demographically diverse cases with a psychotic disorder who underwent comprehensive psychiatric and neurological phenotyping and 115 matched controls. Following genotyping, we calculated polygenic scores (PGSs) for schizophrenia (SZ) and bipolar disorder (BP) using Psychiatric Genomics Consortium GWAS summary statistics. To test if overall genetic risk can be partitioned into affected neurotransmitter pathways, we calculated pathway PGSs (pPGSs) for SZ risk affecting each of four major neurotransmitter systems: glutamate, GABA, dopamine, and serotonin. Psychosis subjects had elevated SZ PGS versus controls; cases with SZ or BP diagnoses had stronger SZ or BP risk, respectively. There was no significant association within psychosis cases between individual symptom measures and overall PGS. However, neurotransmitter-specific pPGSs were moderately associated with specific endophenotypes; notably, glutamate was associated with SZ diagnosis and with deficits in cognitive control during task-based fMRI, while dopamine was associated with global functioning. Finally, unbiased endophenotype-driven clustering identified three diagnostically mixed case groups that separated on primary deficits of positive symptoms, negative symptoms, global functioning, and cognitive control. All clusters showed strong genome-wide risk. Cluster 2, characterized by deficits in cognitive control and negative symptoms, additionally showed specific risk concentrated in glutamatergic and GABAergic pathways. Due to the intensive characterization of our subjects, the present study was limited to a relatively small cohort. As such, results should be followed up with additional research at the population and mechanism level. Our study suggests pathway-based PGS analysis may be a powerful path forward to study genetic mechanisms driving psychiatric endophenotypes.

## Introduction

Psychosis-spectrum disorders, including schizophrenia (SZ), schizoaffective disorder (SA), and bipolar disorder (BP), affect 0.5-2.3% of people worldwide(1–5). Evidence suggests shared etiology for these disorders, supported by family studies and genome-wide association studies (GWASs) showing high interheritability and shared genomic risk(6–8). Given symptom heterogeneity across the psychosis spectrum, a deeper understanding of the biology underlying specific symptoms may drive psychiatry towards improved patient outcomes using stratified medicine.

One approach toward revealing shared neurobiology is identifying transdiagnostic endophenotypes. Recently, the Bipolar & Schizophrenia Network for Intermediate Phenotypes (B-SNIP) Consortium identified three biotypes of psychotic disorders using neuropsychiatric markers, agnostic to diagnoses(9), that were primarily characterized by differences in cognitive control and sensorimotor reactivity. As knowledge of biology underlying symptom heterogeneity expands, reclassifying psychosis patients using biologically grounded phenotypes may allow for more effective, targeted interventions.

Identification of causal and clinically informative genetic components of psychotic disorders has been aided by large-scale GWASs and estimation of overall genetic risk using polygenic scores (PGSs). While large cohorts were initially required to develop PGSs, once defined, research leveraging PGSs in cohorts with phenotyping that extends beyond diagnosis is revealing how genetic burden is associated with specific symptomatology. For example, recent efforts have found associations between SZ PGS and treatment response(10) as well as neurological and cognitive measures(11,12). Such uses of PGS enable study of genetic burden in small cohorts that have been characterized at a level not feasible at the scale required for GWASs focused on the discovery of new risk loci.

While PGS is a useful metric of genetic risk, it fails to assign burden into relevant biological pathways and elucidate mechanisms underlying symptoms. A novel approach to overcome this is pathway-specific PGS (pPGS), which partitions variants into those contained only in genes of a given function or pathway(13). By assigning burden into pathways, it is possible to test how specific processes contribute to phenotype. This has been used to associate miR-137 pPGS with SZ risk(14) and neurological markers(15,16). Another recent implementation identified SZ and BP subjects with risk in pathways targetable by available pharmaceuticals, showing the utility of pPGS in targeted medicine(17). Among pathways relevant to psychosis, glutamate(18–23), GABA(19–24), dopamine(23,25–28), and serotonin(23,28,29) systems have been strongly associated with psychotic disorders and therapeutics.

There is a major opportunity to use pPGS to identify pathway contributions to psychosis symptoms. Here, we investigated the relationship of overall and neurotransmitter-associated PGS to psychotic disorder presentation at the diagnostic and endophenotypic level. We estimated overall and pathway-level PGS for a diverse cohort recruited from California following a first psychotic episode, and tested association between genetic burden, diagnosis, and clinical and neuroimaging endophenotypes. In a cohort of 206 subjects with psychotic disorders versus 115 controls, we found that overall and pathway PGSs were elevated in cases, pPGS was moderately associated with specific endophenotypes, and unbiased clustering on phenotypes showed associations with pPGS and treatment response. Our results demonstrate the power of pPGS to test the relationships between affected biological pathways and transdiagnostic phenotypes.

## Methods

### Study participants

Psychosis subjects were all outpatients within two years of their first psychotic episode. Subjects were selected from an ongoing psychosis research cohort, which includes 196 first-episode SZ-spectrum patients, 53 patients with first-episode BP with psychotic features, and 135 controls aged 12-38. After all quality control (QC), 119 SZ, 39 SA, 48 BP, and 115 controls were included for this study from an ongoing early psychosis research cohort. The study was approved by the University of California, Davis, Institutional Review Board and all subjects gave written consent and were paid for their participation.

### Psychiatric and neuroimaging phenotyping

All participants were assessed using the Structured Clinical Interview for the DSM-IV-TR (SCID I/P)(30). Clinical interviews were conducted by clinicians with masters or doctoral degrees trained to high reliability (kappa > .70; range = .70-1.0). All patients were followed longitudinally and diagnoses were confirmed 6 months after ascertainment. Exclusion criteria for all groups included: Wechsler Abbreviated Scale of Intelligence (WASI) IQ score below 70, alcohol or drug dependence or abuse within 3 months before testing, positive urine toxicology screen for illicit drugs, prior head trauma worse than a Grade I concussion, or contraindication to MRI scanning. Control subjects were excluded for the following additional criteria: any lifetime diagnosis of an Axis I or Axis II disorder or any first-degree relatives with a psychotic disorder. Before testing, a detailed description of the study was provided and written informed consent obtained.

Subjects were evaluated on the Global Assessment of Functioning (GAF)(31), Global Social Functioning scale (GSF)(32), Global Role Functioning scale (GRF)(33), Young Mania Rating Scale (YMRS)(34), Scale for the Assessment of Positive Symptoms (SAPS)(35), Scale for the Assessment of Negative Symptoms (SANS)(35), and Brief Psychiatric Rating Scale (BPRS)(36). Reality distortion, poverty symptoms, and disorganization scores were defined from the BPRS, SAPS, and SANS(37). Treatment response was defined as > 20% decrease in BPRS from baseline(38).

The GSF and GRF were measured at multiple time points; for each scale, we computed an average of the highest and lowest values measured for each subject during the past year and proceeded with these values for subsequent analyses. Finally, we removed item #8 (“Content”) from scores for the YMRS, as this question asks specifically about positive psychotic symptoms and can skew YMRS scores for subjects with schizophrenia. As such, scores on the YMRS used in these analyses more directly represent mania-specific symptoms. At baseline, all patients had BPRS scores ≥5 to offer sufficient resolution to detect a 20% improvement in score at follow-up. For treatment response calculation, BPRS was rescaled to a lowest score of zero (i.e. score of 24=score of 0)(39).

Behavioral and neuroimaging methods are described previously(40). Consequently, we present these methods in a condensed form. The AX-Continuous Performance Task (AX-CPT)(41) was performed during fMRI. In short, the task requires participants to respond to a series of cue and probe letters and correctly identify the target pair (“AX” trials) while correctly rejecting other pairs. The frequency manipulation of trial types creates a prepotent tendency to make a target response when the “X” probe letter is presented. Consequently when a non-A cue is presented and followed by an X (i.e. “BX” trials) the participant must engage proactive control to retain the goal, keep the incorrect cue in mind, and correctly reject the trial at the probe phase. Participants were excluded if performance did not meet a minimal threshold(42). The primary behavioral measure used for this study is d’-context, which represents a contrast of AX hits versus BX false alarms.

Functional Blood Oxygenation Level Dependent (BOLD) data were acquired using a 1.5T GE Signa and 3.0T Siemens TimTrio. Two regions of interest were defined *a priori* and comprised CueB versus CueA contrast, reflecting high versus low cognitive control-related activity. Specifically, bilateral dorsolateral prefrontal cortex (DLPFC) and bilateral superior parietal cortex (SPC) were defined as 5mm radius spheres based on coordinates from two independent datasets(43,44). All fMRI data were preprocessed using SPM8 (Wellcome Dept. of Imaging Neuroscience, London) and included slice timing correction, realignment, normalization to the Montreal Neurological Institute (MNI) template, and smoothing with an 8mm FWHM Gaussian kernel. Individual runs were excluded when framewise displacement measures of movement exceeded 0.45mm (calculated with https://fsl.fmrib.ox.ac.uk/fsl/fslwiki/FSLMotionOutliers) and whole subjects were excluded if more than half of their data exceeded this threshold. All trial types were modeled (CueA, CueB, AX, AY, BX, BY) and correct responses were used to create first-level images.

Based on the exclusion criteria mentioned above, five controls, nine SZ subjects, and two BP subjects were excluded due to excess motion. Two controls, five SZ subjects, and two BP subjects were excluded for poor behavioral performance. Finally, three controls and six SZ subjects were excluded for other reasons, including scanning artifacts, scanner failure, or button pad failure.

### Genotyping, Quality Control, and Imputation

Subjects underwent blood draws using PAXgene blood DNA tubes, which were subsequently stored at −80 °C until DNA extraction. Tubes were thawed at 37 ᵒC for 15 minutes before extracting DNA using the Qiagen QIAamp DNA Blood Mini Kit. The protocol was followed with minor modifications: volumes of blood, protease, lysis buffer, and ethanol were tripled prior to binding DNA to the spin column; the final elution incubation was carried out at 50 ᵒC for five minutes; and DNA was eluted in 100 µl of nuclease-free water. DNA was cleaned on the Zymo Research Genomic DNA Clean & Concentrator-10 kit with minor modifications: an extra two-minute spin in a clean collection tube was added after completing wash steps; samples were incubated at 50 ᵒC for five minutes during elution; and DNA was eluted in 100 µl of nuclease-free water. After cleaning, samples were analyzed on a spectrophotometer and verified to have concentrations ≥ 50 ng/µl and 260/280 and 260/230 ratios ≥ 1.70.

DNA was genotyped using the Illumina (San Diego, California) Infinium PsychArray-24 Kit at the UC Davis DNA Technologies Core. Initial QC was performed using Illumina GenomeStudio following published guidance(45,46). Additional QC was applied using PLINK. First, variants with greater than 5% missingness were removed. We confirmed that no individuals had more than 1% of SNPs missing, nor did we observe any mismatch between genetic sex inferred by PLINK and subjects’ self-reported sex. We removed variants with significantly different missingness rates between cases and controls (p < 0.001) or variants with significant deviation from Hardy-Weinberg equilibrium (p < 1e-6). No severe heterozygosity outliers were observed. A small number of pairs of individuals appeared to be related; in these cases one subject from each pair was randomly excluded from subsequent analyses. Following imputation, the QC filters outlined above were performed again. Additionally, variants with imputation quality INFO scores < 0.7 or empirical INFO scores < 0.8 were removed.

PLINK (v1.9) was used to calculate genetic principal components (PCs) in the 1000 Genomes(47) Phase 3 dataset, which were projected onto our sample. Samples were submitted to the Michigan Imputation Server for genotype imputation, using the full 1000 Genomes(47) Version 3 dataset as the reference panel. Following all QC, 7,608,150 SNPs were available across 338 unrelated subjects. Following additional review, 14 subjects with schizophreniform disorder and 2 subjects with schizotypal disorder that had initially been included were excluded from subsequent analyses due to insufficient numbers, such that the final number of subjects was 321 as described in “Study Participants.”

We additionally checked for presence of known copy number variants (CNVs) of high penetrance for schizophrenia risk in our samples using iPsychCNV5 and PennCNV(48–50), using default parameters. CNVs called by both software were retained as consensus calls for further analysis. This resulted in a total of 333 CNVs across all subjects. We cross-referenced these results with genome-wide significant CNV loci associated with SZ(51) and filtered for CNVs in our sample that overlapped with at least 50% of one of these previously identified SZ risk CNV loci. One subject with SZ appeared to have a previously described SZ risk CNV: 15q11.2 deletion. As the role of deletion at 15q11.2 is currently of uncertain significance in schizophrenia risk(52,53), we retained this individual for all analyses.

### Polygenic Score Calculation

Using Psychiatric Genomics Consortium GWAS summary statistics for SZ (2021)(22) and BP (2022)(54), we employed PRS-CS(55) to calculate SZ and BP PGS, with the phi parameter set to 0.01 as recommended without a validation sample. PRS-CS applies shrinkage to optimize PGS prediction and may not be applicable when restricting PGS to genes from specific pathways. Therefore, we used the PRSet function from PRSice(56) to calculate pPGS for the four neurotransmitter pathways using SZ GWAS summary statistics(22), as our subjects were weighted toward SZ diagnosis and case-control status was best described by SZ PGS. As recommended by the authors of PRSet, in order to avoid eliminating SNPs in certain genes, no p-value filter was applied when calculating pPGS. In order to select the R^2^ cutoff used for clumping, we tested five potential values (0.1, 0.3, 0.5, 0.7, 0.9) in the subset of European ancestry samples. All values returned a significant relationship with case-control status. For results reported here, we used a clumping R^2^ of 0.7 as it showed the strongest association with case status. We report results using alternative clumping values in Fig. S2, which demonstrate stability of our findings across clumping R^2^ values. Otherwise, default parameters of PRSet were retained, with the 1000 Genomes(47) European subset used as the LD reference panel.

To reduce bias from population stratification, we regressed calculated PGSs on the first 10 principal components of genetic variation in our subjects. We then assessed performance of regressed PGSs across different ancestries present in our study sample. Subject ancestries were estimated by assigning each subject to their nearest 1000 Genomes(47) population by minimizing Euclidean distance to population centroid in 20-dimensional PC space, as recommended by Privé et al.(57). Regressed PGSs (hereafter called PGSs) showed substantial reduction in ancestry-specific stratification and appeared to replicate top-level associations in both European/white and non-European/white subjects (Fig. S3).

### pPGS pathways

Pathways include genes relevant in glutamate, GABA, dopamine, and serotonin. Genes were sourced from KEGG(58), REACTOME(59), and AmiGO(60) by searching for pathways and ontologies that include these neurotransmitters or variations on them (e.g. “glutamatergic”). The complete gene list is in Table S1.

### Unbiased clustering

Endophenotypes were z-score normalized within cases. Subjects were clustered on all endophenotypes. We used all subjects with complete endophenotype data (n = 90) to calculate the optimal number of clusters using the NbClust R package(61). Estimates converged on three clusters. Subsequently, we used all subjects with greater than 50% of endophenotype variables available (n = 167) for k-means clustering using the flipCluster R package(62).

### Statistical analysis

All analyses were completed in R(63). One BP subject was excluded as an outlier from analyses based on Cook’s distance near 1 in regression models. Correlation analyses and plots were produced using the psych(64) and corrplot(65) packages. Violin plots were produced using the vioplot package(66). Effect sizes for associations are presented as standardized β coefficients and odds ratios, calculated using the lm.beta(67) R package.

Nagelkerke’s R^2^ was calculated for variance explained in logistic regression models of disease status by PGS using the RMS R package(68). All endophenotypes were tested against PGS and the following covariates in regression models: chromosomal sex, age, self-reported race, self-reported ethnicity, and diagnosis. Protocol/scanner type was controlled for in cognitive control analyses. Overall SZ PGS was included as a covariate for endophenotype analyses by pPGS. Models of continuous variable phenotypes against PGS are represented with partial regression plots.

For cluster versus diagnosis model comparison, we regressed PGSs separately against clusters, plus standard covariates, and against diagnoses, plus covariates. In separate analyses, we regressed PGS against both cluster and diagnosis in the same model, as well as covariates. BP and Cluster 3 were the reference levels for, respectively, diagnosis and cluster.

### Multiple testing corrections

Multiple testing corrections were applied for analyses of phenotype, including diagnosis and case status, by pathway PGS. Given the correlated structure of phenotypes and of pPGS in our subjects, we used the poolr R package(69) to calculate the effective number of statistical tests following Galwey (2009)(70).

This methodology identified effectively five (of initially six) independent PGS variables and 12 (of initially 17) independent endophenotype variables. Total tested dependent variables were multiplied by total tested independent variables such that findings were corrected for 61 tests (five PGS by 12 endophenotypes plus one test for differential treatment response across k-means clusters) using a Benjamini-Hochberg(71) false discovery rate (FDR) of 0.10.

### Permuted null pathway calculation

For each of the pathways in our main analyses, we constructed 10,000 matched null gene sets with an equivalent number of genes, chosen such that the probability of a gene’s inclusion is proportional to its length. We then calculated pPGS for these 40,000 permuted null gene sets in the same way as described above, i.e. using PRSet to calculate pPGS for our target sample and regressing out the first 10 genetic PCs.

## Results

### Demographics

Subjects were demographically heterogenous and relatively well matched across psychosis cases (n = 206) and controls (n = 115). Cases were composed of subjects with SZ, SA, and BP diagnoses (Table 1).

**Table 1:**
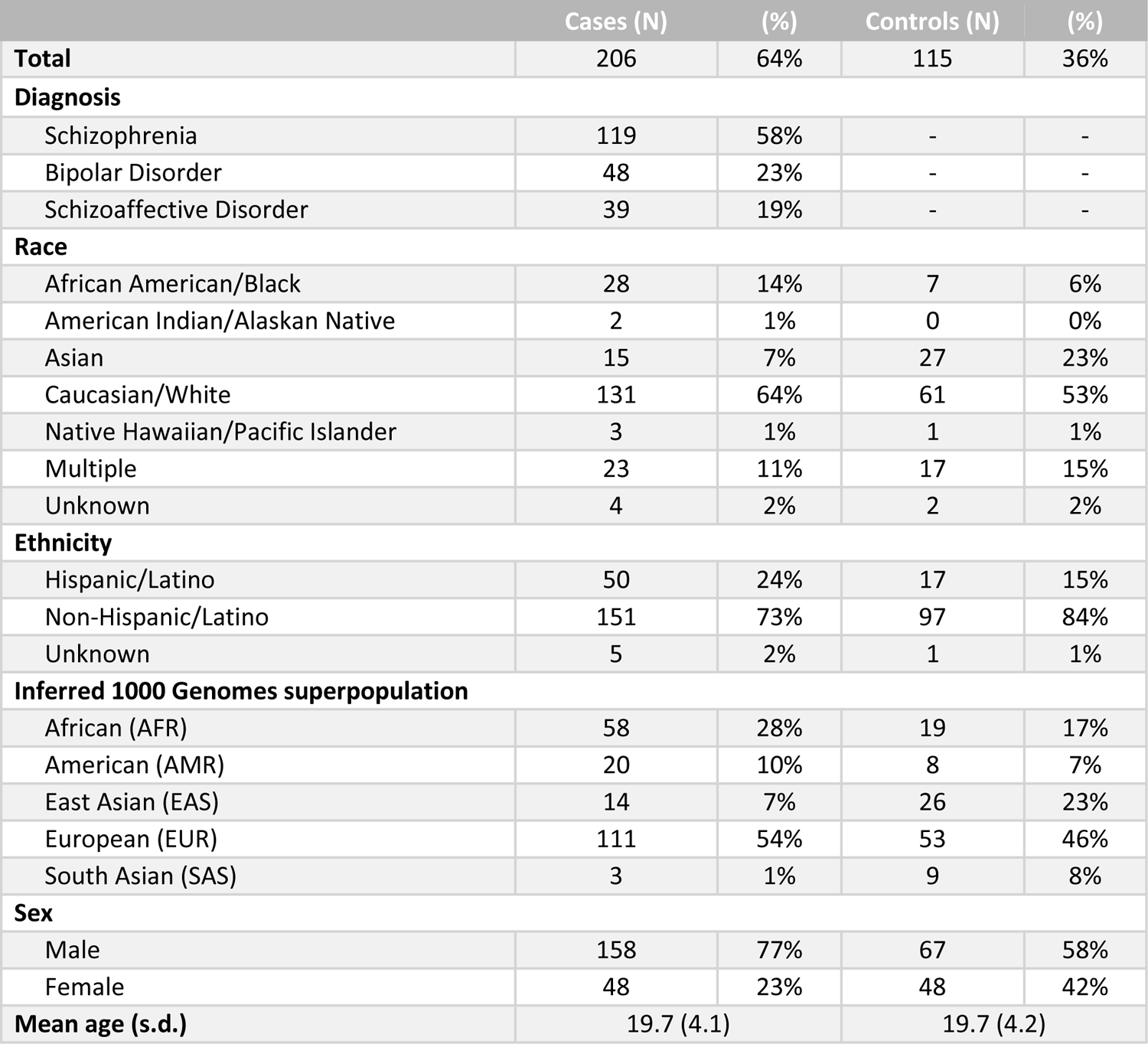
Demographics of study subjects. Study subjects were racially and ethnically diverse across both cases and controls. Potential differences in demographics were considered when controlling for covariates in subsequent analyses. (s.d. = standard deviation)

### Overall and pathway PGSs predict diagnostic status

We first tested whether overall PGS was associated with psychosis status, correcting for latent ancestry differences via inclusion of the top 10 PCs, as well as sex, age, and self-reported race and ethnicity. SZ PGS was associated with case status (OR = 2.69 [CI: 2.06, 3.51]; p = 1.1×10^-4^) (Fig. 1A) and explained 6.3% of variance in status (p = 1.1×10^-4^), similar to previous reports(72–76) (Fig. S1). This association was stronger in SZ (OR = 3.06 [CI: 2.26, 4.14]; p = 2.1×10^-4^) and SA (OR = 3.67 [CI: 2.32, 5.81]; p = 4.7×10^-3^) (Fig. 1B). While BP PGS was not significant in cases overall, it was elevated in BP subjects (OR = 3.55 [CI: 2.34, 5.37]; p = 2.3×10^-3^) (Figs.1A,B). These findings passed multiple testing corrections at FDR < 0.10. SZ and BP PGS were moderately correlated in SZ and SA subjects, but not BP subjects (Fig. 1C). These results were generally consistent across PGS clumping parameters (see Methods, Fig. S2) and when subsetting our cohort to only white European subjects (Fig. S3). Thus, overall PGS captured both transdiagnostic and diagnosis-specific genetic risk for psychotic disorders, consistent with a strong literature on partial genetic overlap between SZ, SA, and BP(6,7,77–79).

**Figure 1:**
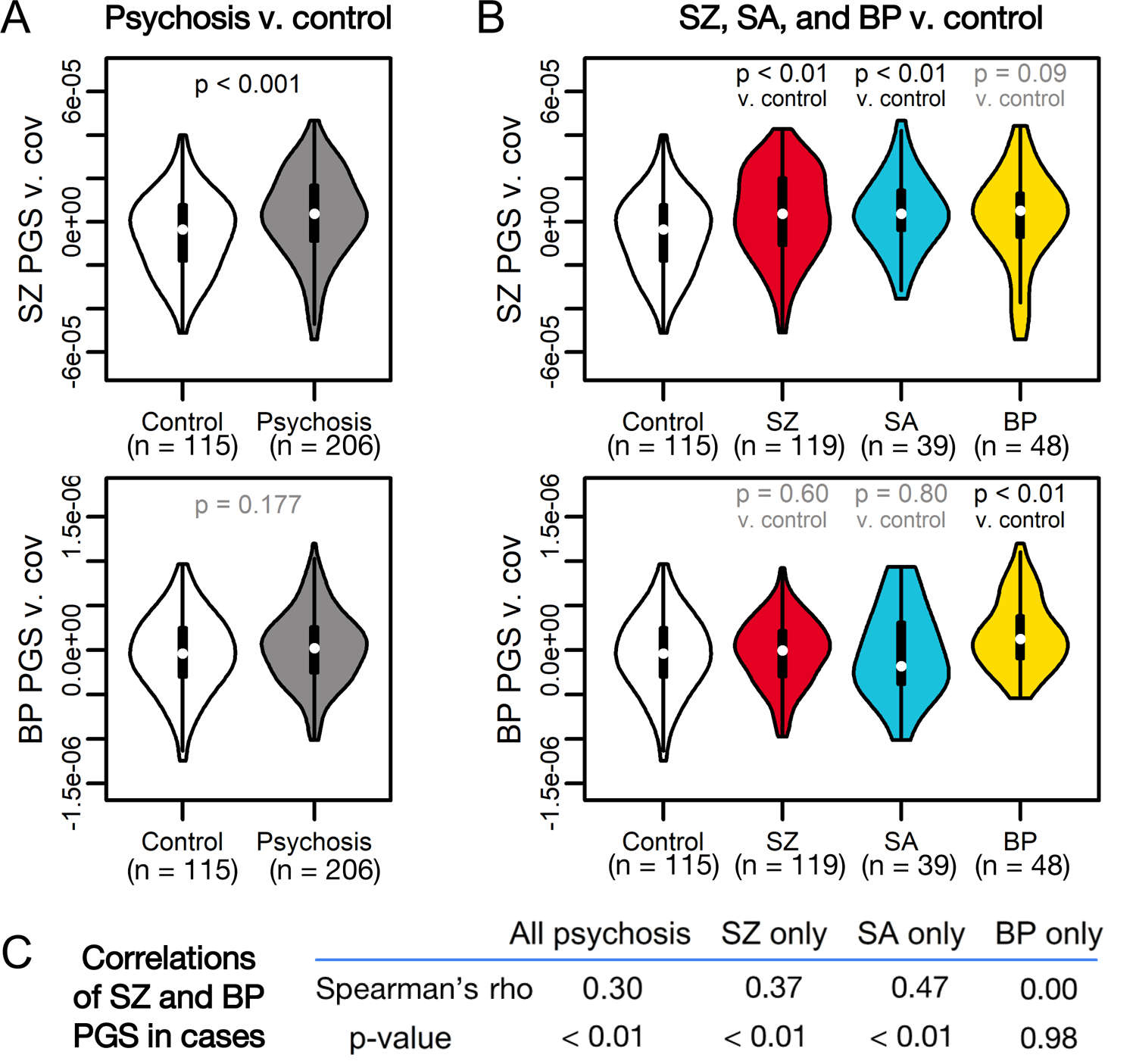
SZ and BP PGS are associated with psychotic disorder diagnoses. (A) SZ PGS (top) is significantly associated with psychosis case status, while BP PGS (bottom) is not. (B) SZ PGS (top) is associated with SZ and SA status versus controls, but not BP status. Conversely, BP PGS (bottom) is associated with BP status versus controls, but not SZ or SA status. For each (A) and (B), the top row represents a comparison of SZ PGS across diagnostic groups, while the bottom row represents a comparison of BP PGS across diagnostic groups. (C) Spearman’s correlations of SZ and BP PGS in psychosis cases show moderate-to-high correlations between PGS for SZ and SA subjects, but not for BP subjects.

### Association of pPGS with phenotypes

As SZ PGS had stronger overall and transdiagnostic association to psychosis in this cohort, we next tested if this risk could be further subset via pPGS associated with major neurotransmitter pathways. To dissect genetic contributions of biological pathways to clinical and neurobiological phenotypes, we calculated pPGS for loci associated with four neurotransmitter systems relevant to psychosis: glutamate(18–22), GABA(19–24), dopamine(25–28), and serotonin(28,29), allowing overlap in genes annotated to each pathway (Fig. 2A, Table S1). These pPGS showed moderate intercorrelations and correlations with overall PGS (Fig. 2B).

**Figure 2:**
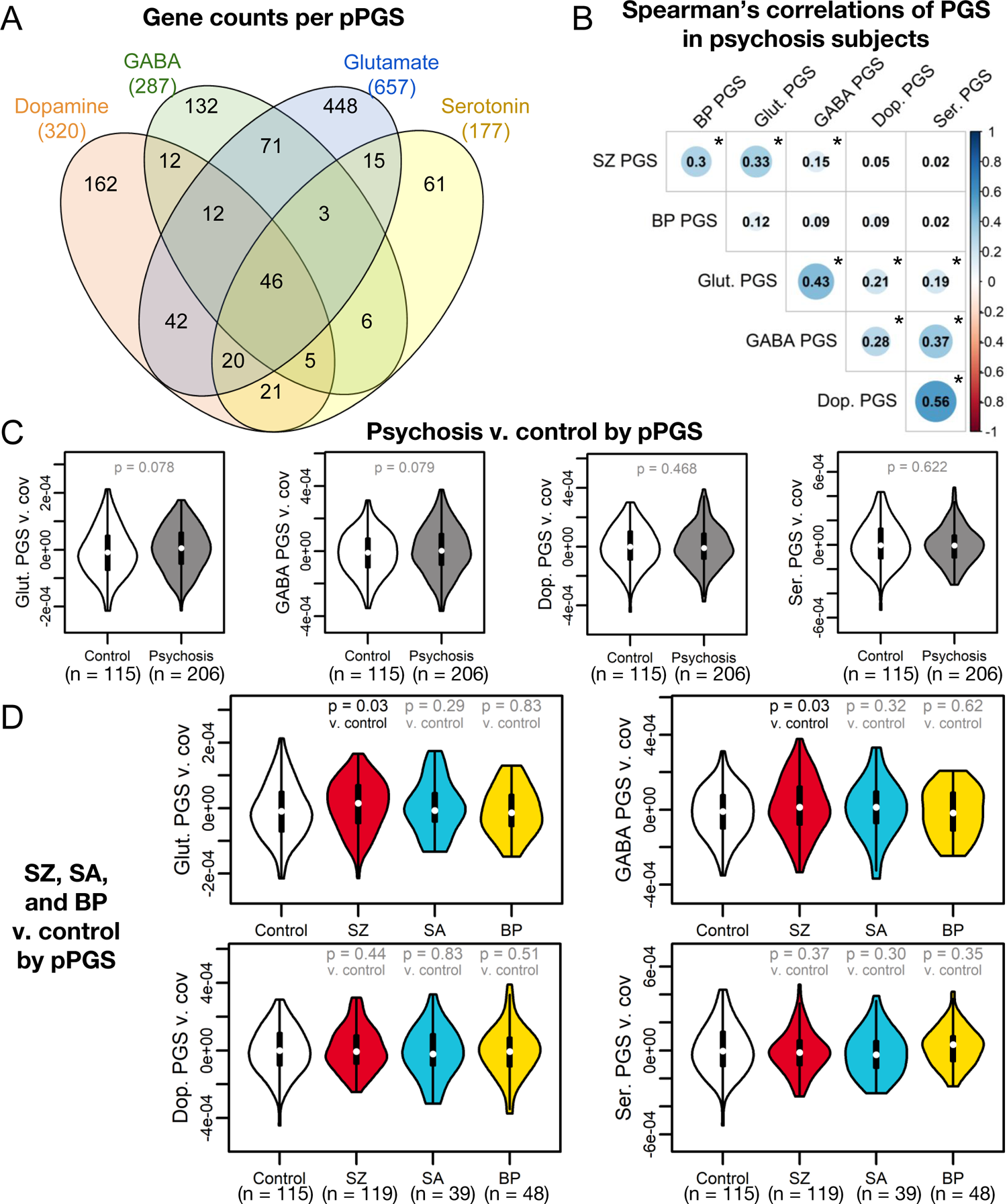
Pathway polygenic scores (pPGS) based on four main neurotransmitter systems. (A) KEGG(58), AmiGO(60), and REACTOME(59) were searched for genes associated with glutamatergic, GABAergic, dopaminergic, and serotonergic neurotransmission. (B) pPGS developed from these gene sets showed substantial correlation in psychosis subjects when regressing against the first four genetic principal components. Statistically significant correlations (p < 0.05) are denoted with an asterisk. (C) While no pPGS significantly separated overall psychosis cases from controls, (D) glutamate and GABA pPGS separated SZ cases specifically from controls, though these findings do not pass FDR < 0.10.

We first tested association with overall disease status. Glutamate and GABA pPGS showed weak trends toward explaining variance in case status (OR = 1.57 [CI: 1.22, 2.04] to 1.58 [CI: 1.22, 2.05]; both p = 0.08; Fig. 2C). Glut and GABA pPGS showed improved explanation of SZ status over a null background distribution of size-matched gene sets (Fig. S4) and were modestly elevated in SZ subjects relative to controls (OR = 1.90 [CI: 1.42, 2.56] to 1.92 [CI: 1.43, 2.57]; both p = 0.03; Fig. 2D), though case-control separation did not pass multiple testing corrections. This suggests disease status in our cohort is partially explained by glutamatergic and GABAergic genetic risk.

### pPGSs are associated with endophenotypes in cases

We next tested if PGS was associated with symptom variation across 10 phenotypes within subjects with a psychotic disorder (Fig. 3A). For these analyses, we tested for associations only within psychosis cases, as data on psychosis endophenotypes was only collected for subjects with psychosis. Overall PGSs were not significantly associated with measured phenotypes (Fig. S5). However, we found evidence for associations of pathway-specific risk to six phenotypes in global functioning, role functioning, cognitive control, and treatment response, though only DLPFC β-values by glutamate and GAF by dopamine passed multiple testing corrections at FDR < 0.10. Full data on non-significant relationships is available in Fig. S5.

**Figure 3:**
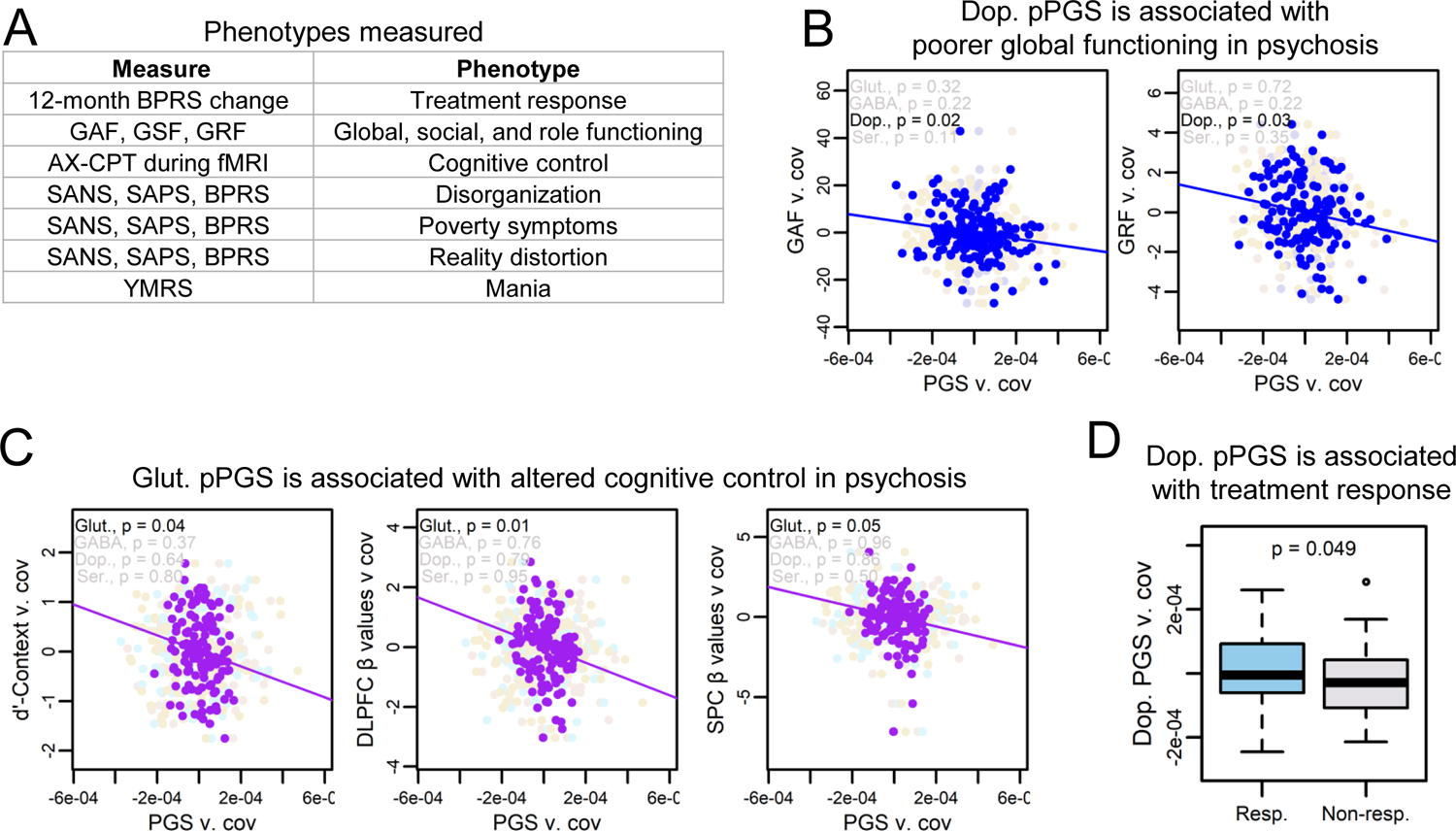
pPGSs identify mechanisms for endophenotypes of psychotic disorders. (A) Subjects were phenotyped on a range of psychological, clinical, and neurological measures. (B) Dopamine pPGS was associated with poorer global functioning and role functioning in psychosis subjects when controlling for diagnosis and overall SZ PGS. Only the association with global functioning passed FDR < 0.10. (C) Glutamate pPGS showed associations with poorer performance and reduced cortical activation in psychosis subjects during the AX-CPT cognitive control task when controlling for diagnosis and overall SZ PGS. Only the association with DLPFC β-values passed FDR < 0.10. (D) Treatment response showed a slight association with increased dopamine pPGS, though this did not pass FDR < 0.10. (Resp. = responder to treatment, non-resp. = non-responder to treatment)

Dopamine pPGS was associated with poorer global (ꞵ = −0.17 [CI: −0.25, −0.10]; p = 0.02) and role (ꞵ = −0.19 [CI: −0.28, −0.10]; p = 0.03) functioning, though role functioning did not pass multiple testing corrections (Fig. 3B). Perhaps the strongest finding was an association between glutamate pPGS and cognitive control in cases, though only the association to DLPFC ꞵ-values passed multiple testing corrections (DLPFC ꞵ-values: ꞵ = −0.22 [CI: −0.31, −0.13]; p = 0.01) (SPC ꞵ-values: ꞵ = −0.19 [CI: −0.29, −0.09]; p = 0.05) (d’-Context: ꞵ = −0.18 [CI: −0.26, −0.09]; p = 0.04) (Fig. 3C). This finding is consistent with a prominent hypothesis in schizophrenia literature that perturbations to glutamatergic signaling may be associated with deficits in cognitive control(80–83). Notably, our findings represent results from a relatively small cohort and should be followed up in larger cohorts and with more detailed mechanistic studies, and only one endophenotype association to glutamate passed multiple testing corrections.

However, these results show the promise of using pPGS to partition genome-wide risk into hypothesized mechanistic pathways toward symptoms of psychiatric disease. Of 63 subjects with available treatment response data, 35 were responders, consistent with reported efficacy rates(84). Though underpowered, treatment response showed a weak association with higher dopamine pPGS which did not pass multiple testing corrections (OR = 3.40 [CI: 1.83, 6.34]; p = 0.05) (Fig. 3D). While this finding should be considered preliminary, this is consistent with literature associating dopamine with antipsychotic efficacy.

### Unbiased phenotype clustering of psychosis cases and genetic burden

Biotype-level grouping of psychosis cases, such as by B-SNIP(9), may advance the use of biological data to better model psychopathology. We hypothesized a biotype-style approach might unmask explanations of psychosis symptom heterogeneity by overall and pathway-level PGS. Toward this goal, we used k-means clustering to group endophenotype presentation among cases. We clustered subjects on z-score normalized phenotype data into three groups following cluster optimization using the NbClust R package(61).

Clusters showed distinct symptom profiles (Fig. 4A, Table S2). Cluster 1 was distinguished by high mania, disorganization, and reality distortion, and moderate impairments in cognitive control. Cluster 2 had deficits in negative symptoms, showed poor outcomes in role and social functioning, and had the poorest measures of cognitive control. Cluster 3 had low pathology. Cluster 1 showed evidence of enrichment for treatment responders relative to Clusters 2 and 3, though this did not reach significance (p = 0.057) (Fig. 4B). In contrast, treatment response showed no separation by diagnosis (p = 0.845).

**Figure 4:**
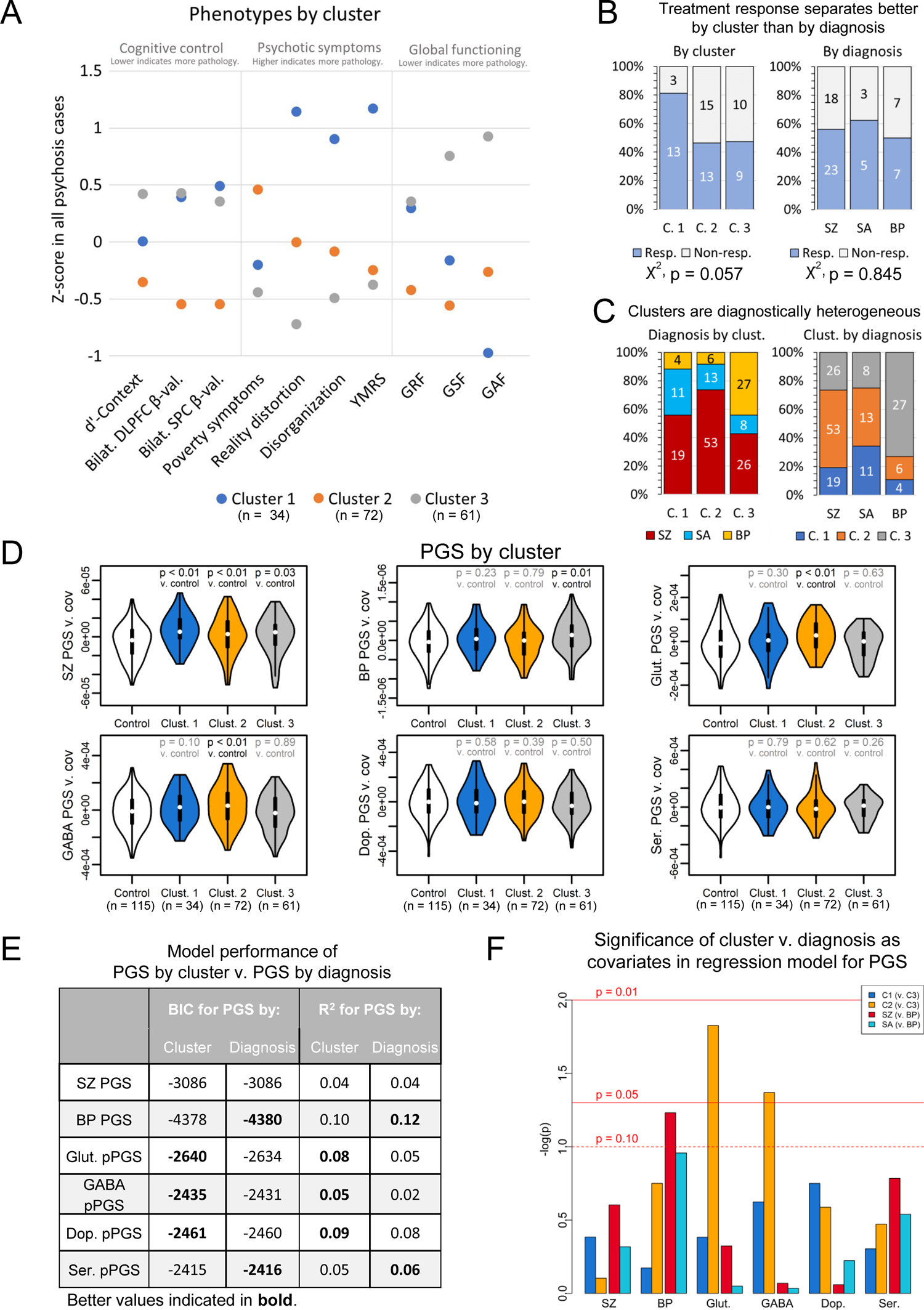
Psychosis subjects separate into clusters based on symptom profiles. (A) Psychosis subjects separate into three clusters that are characterized by (Cluster 1) mania, disorganization, reality distortion, and moderate impairments in cognitive control; (Cluster 2) high poverty symptoms, poor social functioning, and strong deficits in cognitive control; and (Cluster 3) overall low levels of pathology. (B) Cluster 1 showed enrichment for treatment responders, though this did not pass FDR < 0.10. In contrast, treatment efficacy did not separate by diagnosis. (C) Diagnoses are mixed across clusters. (D) SZ PGS is elevated in all clusters, though this did not pass FDR < 0.10 for Cluster 3. BP PGS is elevated in Cluster 3. Glutamate and GABA pPGS are elevated in Cluster 2. (E) Models regressing PGS against cluster perform better for glutamate and GABA pPGS than models regressing PGS against diagnosis. Models regressing PGS by diagnosis perform better than PGS by cluster for BP PGS. Weaker evidence for performance differences by model type are present for dopamine pPGS and serotonin pPGS. (F) Glutamate and GABA pPGS are significantly associated with cluster membership when controlling for diagnosis. (C. 1 = Cluster 1, C. 2 = Cluster 2, C. 3 = Cluster 3, resp. = responder to treatment, non-resp. = non-responder to treatment)

Clusters were demographically and diagnostically heterogenous (Fig. 4C, Table S2), though Cluster 3 captured most BP subjects. By contrast, SZ and SA subjects were relatively evenly distributed across Clusters 1 and 2. This suggests that, among endophenotypes available in our subjects, Clusters 1 and 2 captured relatively strong endophenotype differences that are transdiagnostic across SZ and SA subjects.

We next tested associations between cluster and PGS (Fig. 4D, Table S2). SZ PGS was associated with all clusters, though this did not pass multiple testing corrections for Cluster 3 (Clust. 1: OR = 4.90 [CI: 3.01, 7.96]; p = 1.1×10^-3^) (Clust. 2: OR = 2.59 [CI: 1.83, 3.66]; p = 6.1×10^-3^) (Clust. 3: OR = 2.21 [CI: 1.53, 3.19]; p = 0.03). BP PGS was strongly associated with Cluster 3 (OR = 2.51 [CI: 1.74, 3.60]; p = 0.01), consistent with its preponderance of BP subjects. Cluster 3 was not associated with any pPGS, suggesting genetic risk may lie outside of SZ variants in neurotransmitter-linked loci. In contrast, GABA (OR = 2.51 [CI: 1.78, 3.53]; p = 0.01) and glutamate (OR = 3.21 [CI: 2.24, 4.59]; p = 1.15×10^-3^) pPGS were elevated in Cluster 2, both of which passed multiple testing corrections. This may have implications for the symptom profiles of this cluster. Cluster 2 showed strong negative symptoms, which are thought to largely arise from glutamatergic dysfunction(85,86). Cluster 2 also showed deficits in social and role functioning, which are predicted by negative symptoms(87), and strong cognitive control deficits. pPGS findings suggest Cluster 2’s symptoms in our cohort may be more specific to GABAergic and glutamatergic risk, consistent with the hypothesis of an imbalance between these neurotransmitters in psychosis(20). Following up on these findings in larger cohorts may eventually prove useful in identifying pathways underlying transdiagnostic psychiatric symptoms.

We next compared explanatory power of our clusters versus diagnoses to model genetic risk in our cohort. For this, we defined models regressing PGSs against either cluster or diagnosis. Per Bayesian information criteria and R^2^ (Fig. 4E), models using clusters performed better than models using diagnosis to predict, in particular, glutamatergic and GABAergic pPGS. We next regressed PGS against both cluster and diagnosis as covariates to test independence between diagnosis and cluster (Fig. 4F). Again, glutamate (ꞵ = 0.23 [CI: 0.14, 0.33]; p = 0.02) and GABA (ꞵ = 0.20 [CI: 0.10, 0.29]; p = 0.04) pPGS were associated with Cluster 2 when including diagnosis in the model. These results demonstrate that, within subjects of the same diagnosis, endophenotypes captured here showed specific associations to glutamatergic and GABAergic pPGS. These results provide evidence for the biological validity of these clusters overall within our cohort and within the context of molecular pathways.

## Discussion

We tested how overall and pathway-specific PGS are related to diagnosis and variation in psychosis symptoms in a diverse patient sample. We calculated overall SZ and BP PGS, and further curated gene sets representing four major neurotransmitter systems(58–60), comparing pPGS for SZ variants in these pathways. Overall genome-wide PGS, and to a weaker degree glutamate and GABA pPGS, were associated with case status, though glutamate and GABA did not pass multiple testing corrections. pPGSs outperformed overall PGS in explaining endophenotypes. Dopamine was significantly associated with poorer overall functioning and marginally associated with treatment response, though the association to treatment response did not pass multiple testing corrections. This is a preliminary but intriguing result, suggesting the hypothesis for future research that patients with increased dopaminergic risk may be better candidates for antipsychotic medications. Our results also showed associations between glutamate pPGS and cognitive control. Unsupervised clustering identified three phenotypically distinct groups of psychosis subjects that differed predominantly on positive symptoms, negative symptoms, cognitive control, and global functioning. Though our data largely represent different phenotypic measurements than those used by B-SNIP, we reproduced those findings which our measures can capture and generated preliminary hypotheses on the relationship between biotypes and genetic burden. Like B-SNIP, we identified three diagnostically mixed endophenotypes in our subjects that separate on general impairment and cognitive control. Notably, Cluster 2, which showed primary deficits in cognitive control, negative symptoms, and social functioning, had elevated glutamatergic and GABAergic risk. Treatment efficacy showed some evidence for separation by cluster. Within diagnoses, clustering on available endophenotypes unmasked different symptom presentations that showed specific profiles of genetic risk. Our results require replication before generalization outside of our small cohort, but may serve as proof-of-principle for the use of biologically partitioned polygenic risk to study shared mechanisms behind psychiatric symptoms.

Our study features limitations. Notably, our cohort is smaller than many genomics studies, though our deep level of phenotyping is a strength and generally not possible for the larger or combined cohorts needed for GWAS. As a result of our small sample, our results should be considered preliminary data and are best understood as a model for the use of pPGS to study hypothesized mechanisms of disease. Additional cohorts featuring endophenotypes not captured in our study, such as mood symptoms or potential biomarkers of disease, may help draw a more complete picture of how genetic burden drives psychiatric symptomatology. Focusing on PGS rather than SNP discovery mitigates the limitations of our sample size, though limits our study to integration of previously-defined associations. We tested *a priori*-defined and hypothesis-driven pathways in the present study, however, there are certain to be other contributing pathways not investigated here. While diverse cohorts are needed to capture generalized genetic risk across populations, there is currently no standard for transposing PGS across ancestries.

Here, we regressed PGS on the first 10 genetic PCs and additionally controlled for race and ancestry in association analyses and show that our methods generally address ancestry differences in PGS in our cohort. However, future studies on larger diverse replication cohorts will be needed. Finally, k-means clustering will identify clusters even when the true nature of the data is continuous or cluster differences are not biologically relevant. The identification of unique PGS profiles for our clusters, as well as evidence for by-cluster differences in treatment response, provide auxiliary evidence supporting separate mechanisms driving distinct clusters of primary symptoms within our subjects. PGSs showed associations to specific clusters even within subjects of the same diagnosis. Additionally, we replicate key findings from previous clustering attempts on psychosis subjects(9).

Our results show the utility of overall and pathway-specific PGS to understand genetic burden and interrogate convergent neurobiological mechanisms underlying disease phenotypes. The neurotransmitter systems tested here have been robustly associated with psychotic disorders(18–29). However, their relationship to specific symptoms have not been elucidated. Mechanisms underlying cognitive and negative symptoms in psychosis are particularly poorly understood, despite being some of the strongest predictors of poor outcomes in psychotic disorders(88–91). Our results support a role for glutamate dysfunction in cognitive, as well as negative and social, symptoms in psychosis. This adds to a wide literature hypothesizing that the biological basis of these symptoms may be partly explained by genetic risk driving glutamatergic abnormalities. Our study and findings represent a model for going beyond overall genetic burden in investigation of the genetic components of psychosis, and, in particular, of maximizing power in smaller but deeply characterized cohorts. While our findings will need to be replicated, they offer a promising look into how genetic burden can be partitioned and can explain specific psychosis diagnoses and endophenotypes. Our work is a step towards a more precise understanding of convergent molecular mechanisms underlying psychotic symptoms, which is critical for realizing the promise of stratified medicine and targeted treatments for those suffering from psychotic disorders.

## Data Availability

All data produced in the present study are available upon reasonable request to the authors.

## Acknowledgements

Subject recruitment, blood draws, and phenotyping were supported by NIMH grant R01MH059883. DNA extraction and genotyping were supported by the UC Davis-HKU Collaboration Grant. Analyses and writing were supported by the UC Davis LaMP T32 (NIMH T32MH112507), the NIMH NRSA F31MH129135, and NIGMS grant R35GM119831.

## Conflict of interest statement

The authors declare no conflicts of interest or competing financial interests.

## Notes

### Competing Interest Statement

The authors have declared no competing interest.

### Funding Statement

This study was funded by NIMH grant R01MH059883, the UC Davis-HKU Collaboration Grant, the UC Davis LaMP T32 (NIMH T32MH112507), the NIMH NRSA F31MH129135, and NIGMS R35GM119831.

### Author Declarations

The IRB of UC Davis gave ethical approval for this work.

### Summary of Updates

Updated PGS calculations and modeling, improved controls for demographics and multiple testing burden.

