## Supplementary Material for "Association of neurotransmitter pathway polygenic risk with specific symptom profiles in psychosis"

### Supplementary figures and tables

**Fig. S1:** Explanatory power of genome-wide PGS for disease status in our cohort is comparable to that previously reported in the literature.

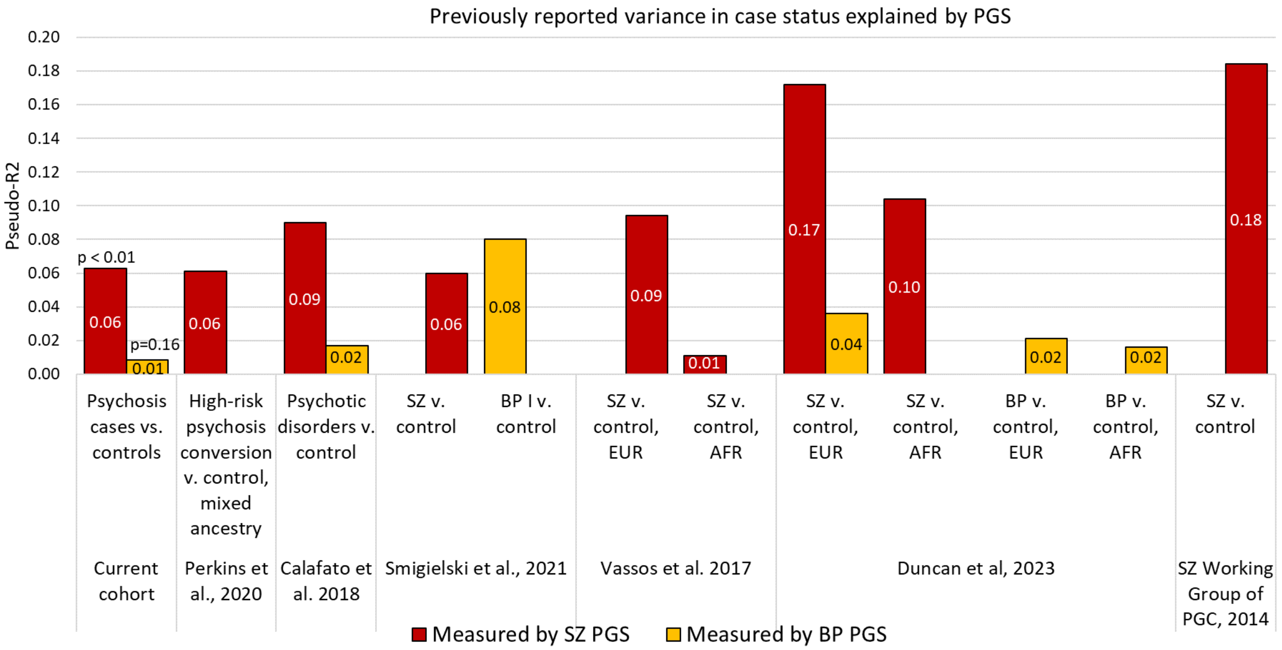

**Fig. S2:** Significant findings at a clumping LD $R^2$ of 0.7 show moderate stability at other $R^2$ values.

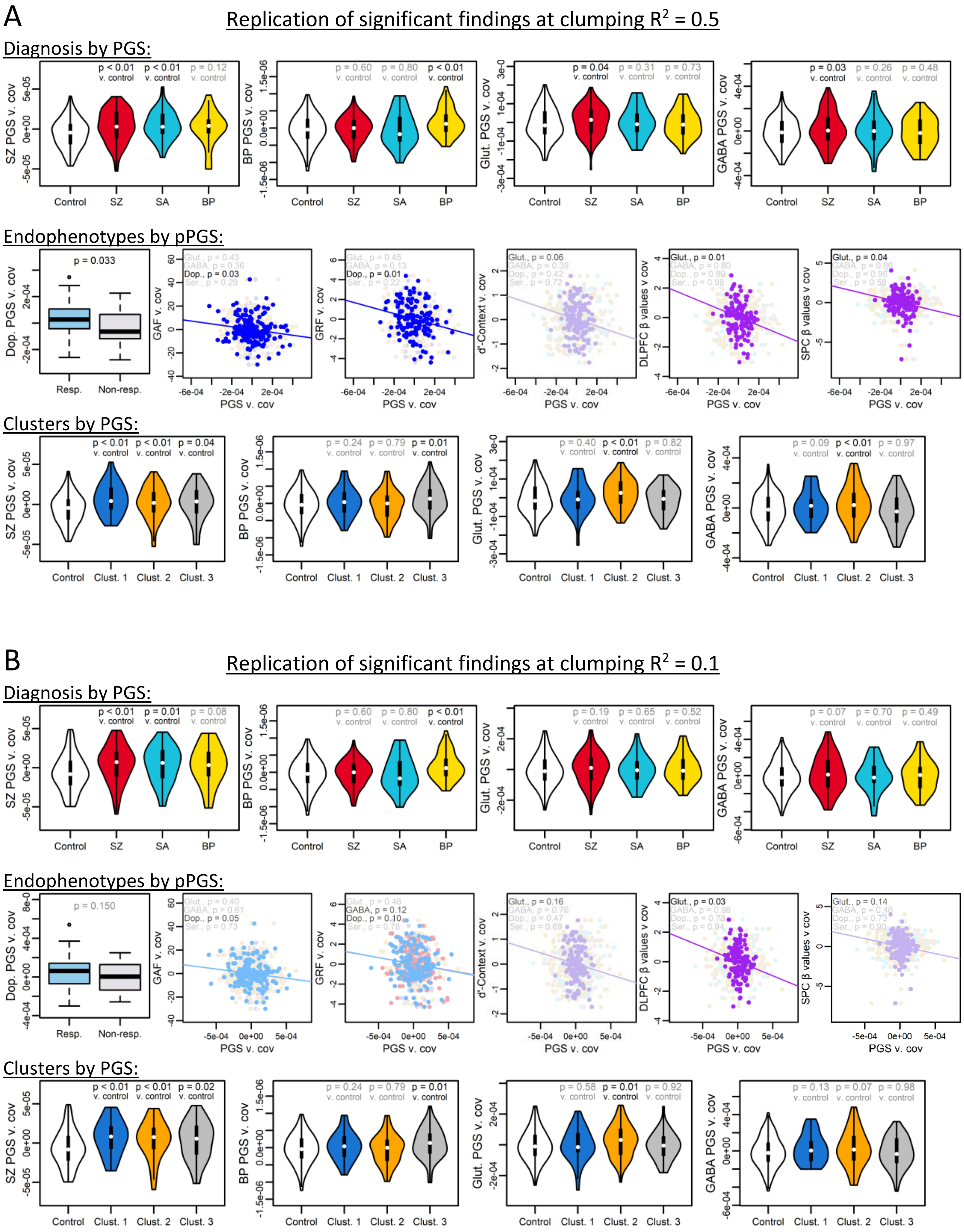

C

Replication summary

| Finding | p at $R^2$ = 0.7 | p at $R^2$ = 0.5 | p at $R^2$ = 0.1 |
|---|---|---|---|
| SZ vs. control by SZ PGS | < 0.01 | **< 0.01** | **< 0.01** |
| SA vs. control by SZ PGS | 0.01 | **< 0.01** | **0.01** |
| BP vs. control by BP PGS | < 0.01 | **< 0.01** | **< 0.01** |
| SZ vs. control by glutamate pPGS | 0.03 | **0.04** | 0.20 |
| SZ vs. control by GABA pPGS | 0.03 | **0.03** | *0.07* |
| Treatment response by dopamine pPGS | 0.05 | **0.03** | 0.15 |
| GAF by dopamine pPGS | 0.02 | **0.01** | *0.05* |
| GRF by dopamine pPGS | 0.03 | **0.01** | 0.10 |
| d'-Context by glutamate pPGS | 0.04 | **0.06** | 0.16 |
| DLPFC β-value by glutamate pPGS | 0.02 | **0.01** | **0.03** |
| SPC β-value by glutamate PGS | 0.05 | **0.04** | 0.14 |
| Clust. 1 vs. control by SZ PGS | < 0.01 | **< 0.01** | **< 0.01** |
| Clust. 2 vs. control by SZ PGS | < 0.01 | **< 0.01** | **< 0.01** |
| Clust. 3 vs. control by SZ PGS | 0.03 | **0.04** | **0.02** |
| Clust. 3 vs. control by BP PGS | 0.01 | **0.01** | **0.01** |
| Clust. 2 vs. control by glutamate pPGS | < 0.01 | **< 0.01** | **0.01** |
| Clust. 2 vs. control by GABA pPGS | < 0.01 | **0.01** | 0.07 |

P-values in **bold text** show strong evidence of replication (less than p ± 0.02 from original result).
P-values in *italic text* show moderate evidence of replication (less than p ± 0.05 from original result).
P-values in blue text show weak evidence of replication (less than p ± 0.10 from original result).
P-values in gray text show little or no evidence of replication (greater than p ± 0.10 from original result).

**Fig. S3:** Regression against 10 PCs effectively normalizes PGS performance across ancestries in the present cohort. (A) Raw SZ PGS shows strong differences across ancestries, while (B) SZ PGS shows no separation by ancestry within-cases or within-controls after regression. (C) Though underpowered due to additional subsetting of our cohort, main findings show evidence of replication when analyses are constrained to only white/non-Hispanic subjects with estimated EUR ancestry. (Estimated ancestries abbreviated as follows: AFR = African, AMR = American, EAS = East Asian, EUR = European, SAS = South Asian) (The y-axes of plots A and B are matched for range to improve comparability.)

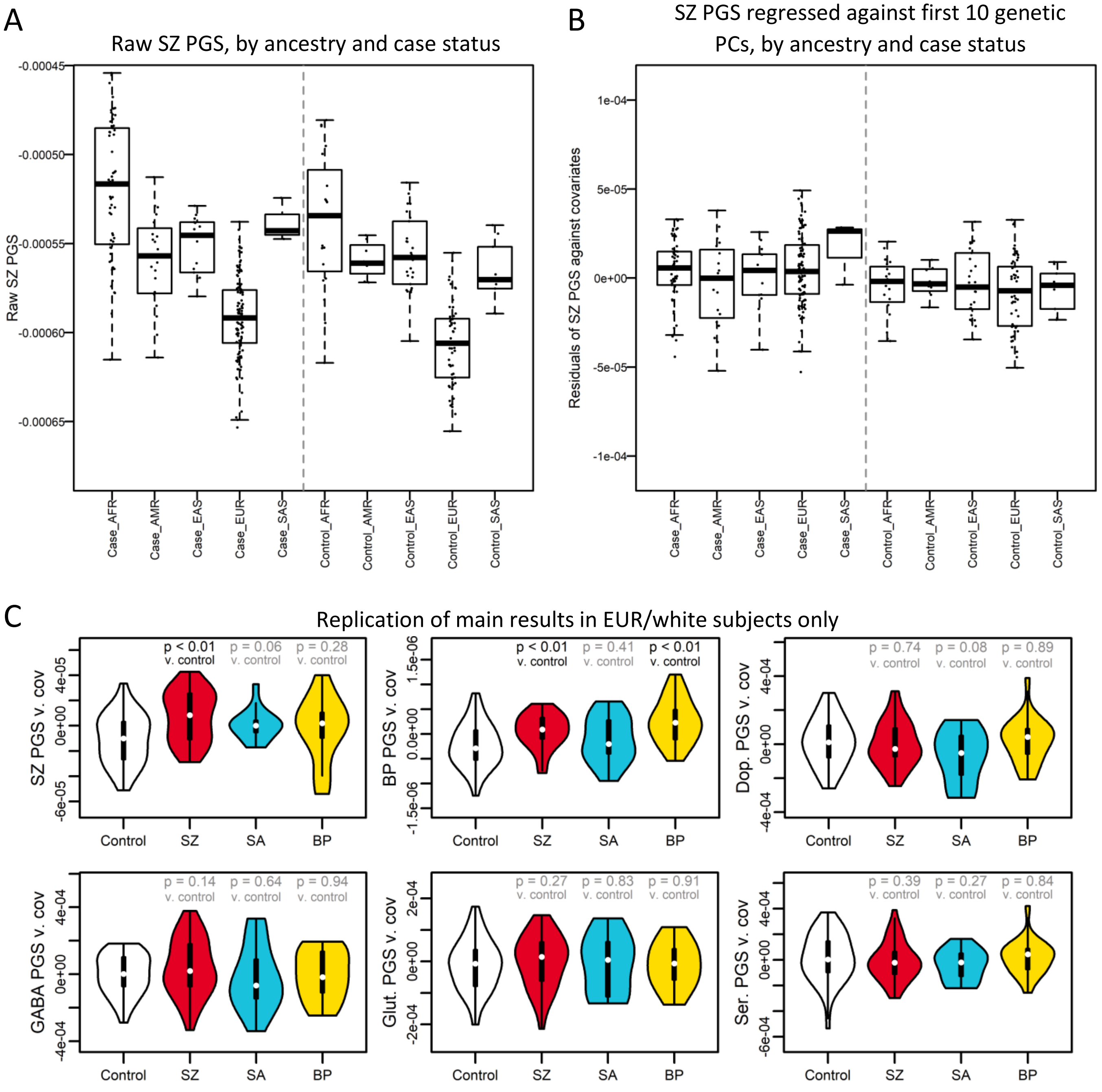

**Fig. S4:** Glutamate and GABA pPGSs show a weak trend toward partially explaining case status in SZ vs. control subjects, and show evidence of increase over a null background of 10,000 size-matched random gene sets. Dopamine and serotonin PGS show no explanatory power for case vs. control status. Red bar indicates Nagelkerke's $R^2$ for the neurotransmitter pPGS. pPGSs are plotted against their respective null distributions.

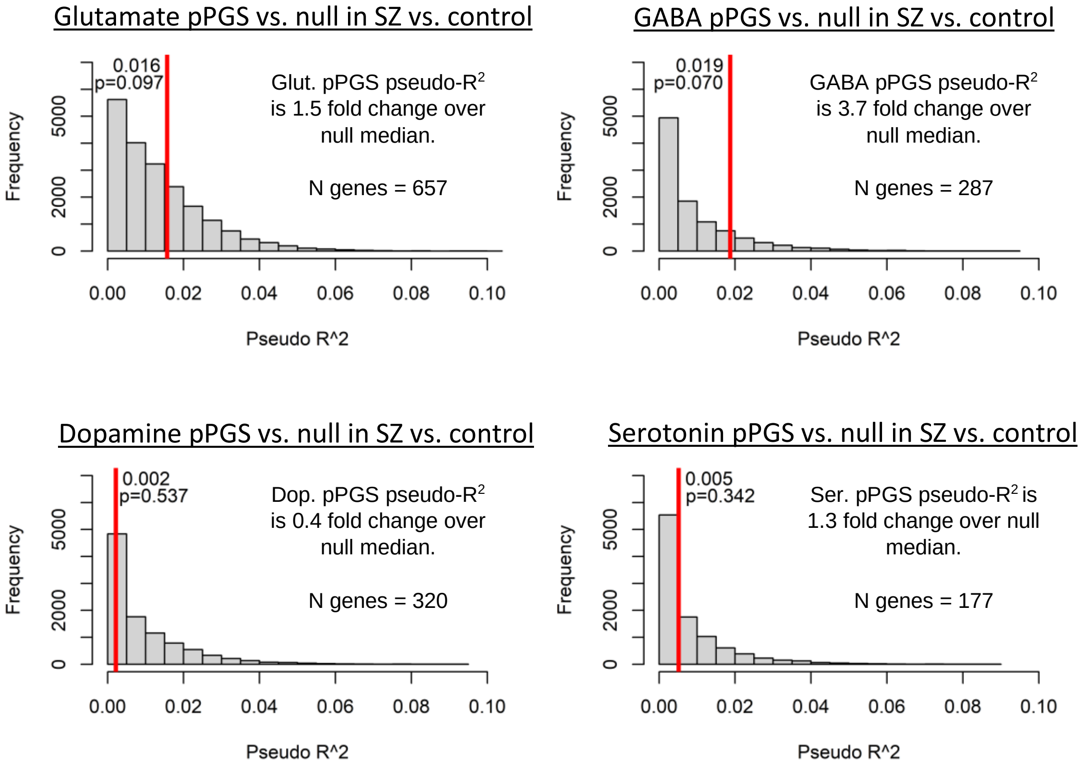

**Fig. S5:** (A,B) Phenotypes measured in this study were not significantly associated with overall (A) SZ PGS or (B) BP PGS in our cohort. (C) Non-significant relationships between pPGS and endophenotypes.

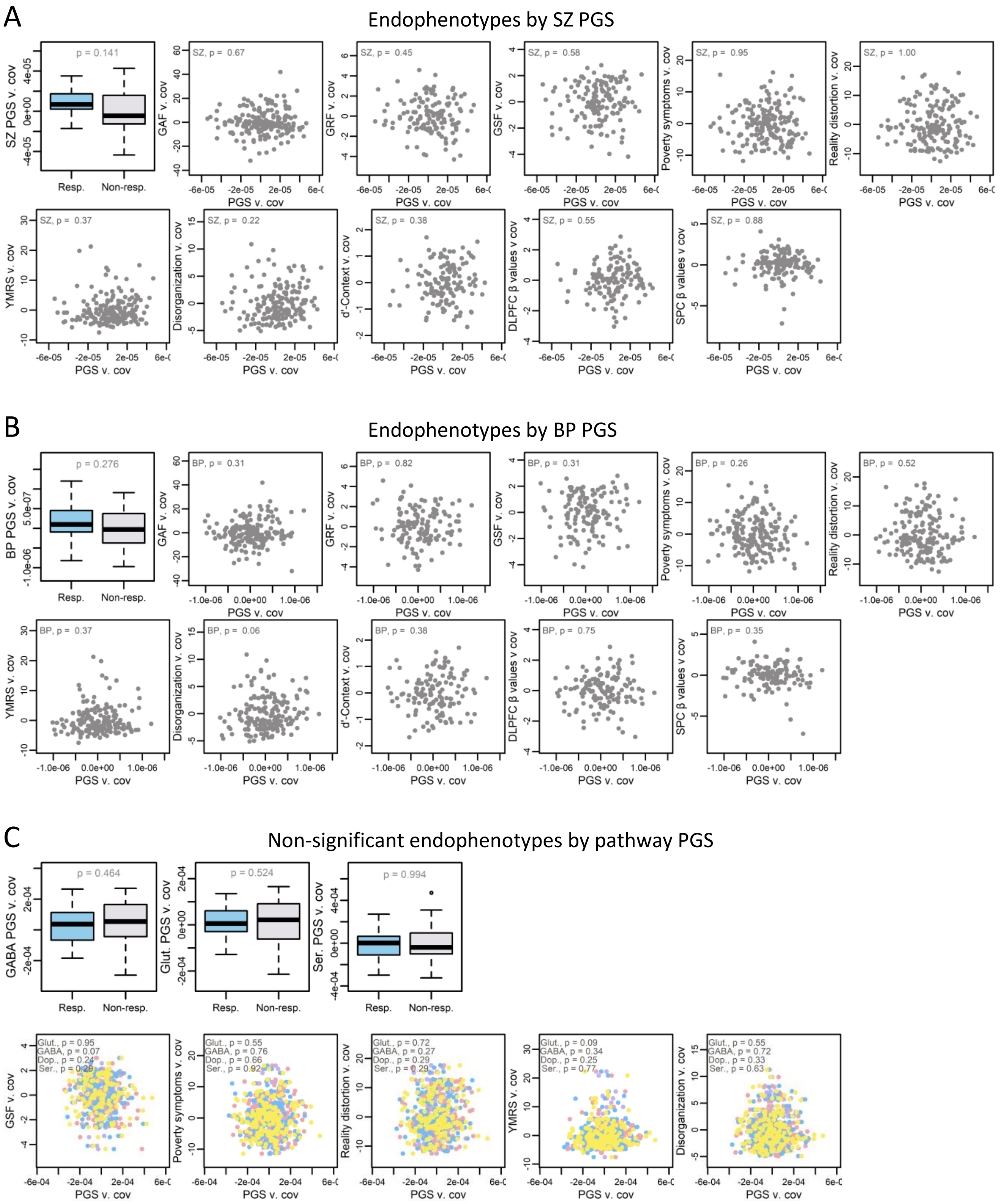

**Table S1:** Genes in neurotransmitter pathways
Data is available as a downloadable Excel file.

**Table S2:** Summary of k-means clusters.

| | Cluster 1 | Cluster 2 | Cluster 3 |
|---|---|---|---|
| **N** | 34 | 72 | 61 |
| **Diagnoses** | SZ: 19 (56%)<br>SA: 11 (32%)<br>BP: 4 (12%) | SZ: 53 (74%)<br>SA: 13 (18%)<br>BP: 6 (8%) | SZ: 26 (43%)<br>SA: 8 (13%)<br>BP: 27 (44%) |
| **Primary symptoms** | ▸ Reality distortion<br>▸ Disorganization<br>▸ Mania<br>▸ Social functioning<br>▸ Global functioning | ▸ Cog. control<br>▸ Poverty symptoms<br>▸ Role functioning<br>▸ Social functioning<br>▸ Global functioning | (mild deficits) |
| **Highest PGS** | ▸ SZ PGS | ▸ SZ PGS<br>▸ Glutamate PGS | ▸ BP PGS |
| **Race** | | | |
| African American/Black | 3 (9%) | 13 (18%) | 6 (10%) |
| American Indian/Alaskan Native | 1 (3%) | 0 (0%) | 1 (2%) |
| Asian | 2 (6%) | 8 (11%) | 3 (5%) |
| Caucasian/White | 24 (71%) | 42 (58%) | 41 (67%) |
| Native Hawaiian/Pacific Islander | 0 (0%) | 2 (3%) | 1 (2%) |
| Multiple/unknown | 4 (12%) | 7 (10%) | 9 (15%) |
| **Ethnicity** | | | |
| Hispanic/Latino | 10 (29%) | 16 (22%) | 46 (75%) |
| Non-Hispanic/Latino | 24 (71%) | 55 (76%) | 14 (23%) |
| Unknown | 0 (0%) | 1 (1%) | 1 (2%) |
| **Inferred 1000 Genomes SuperPopulation** | | | |
| AFR | 8 (24%) | 25 (35%) | 12 (20%) |
| AMR | 5 (15%) | 7 (10%) | 5 (8%) |
| EAS | 1 (3%) | 6 (8%) | 5 (8%) |
| EUR | 20 (59%) | 32 (44%) | 39 (64%) |
| SAS | 0 (0%) | 2 (3%) | 0 (0%) |
| **Sex** | | | |
| Male | 28 (82%) | 59 (82%) | 45 (74%) |
| Female | 6 (18%) | 13 (18%) | 16 (26%) |
| **Mean age (s.d.)** | 20.4 (4.3) | 19.1 (4.3) | 20.0 (4.0) |
